# Effects of anesthesia and oral cleft types on academic achievement

**DOI:** 10.1101/2020.05.21.20108902

**Authors:** Saskia Gladys Nadal

## Abstract

**Objective:** Research looking simultaneously at the effects of anesthesia and oral cleft types on academic achievement is scarce. Available results are contradictory: some studies suggest that anesthesia exposure is responsible for underachievement, and others that responsibility falls instead on the type of orofacial cleft. This study investigates the potential compound effects of exposure to anesthesia and orofacial cleft types on the risk of academic underachievement.

**Design:** Centre Labio Palatin Albert Coninck, Cliniques universitaires Saint-Luc, Brussels, Belgium, nationwide register-based study.

**Setting:** Birth cohort 1995-2007.

**Patients:** Twenty-nine children with isolated orofacial clefts exposed to anesthesia.

**Interventions:** Average duration of exposure to anesthesia before the Certificat d Etudes de Base (CEB) exam was 382 minutes.

**Main Outcome Measure(s):** Scores obtained by patients at the CEB exam were compared with the scores of the 6th grade Belgian general population who passed the same exam controlling for gender, school year, year they passed the exam, medical illnesses, duration of exposure to anesthesia in minutes and socioeconomic confounders.

**Results:** Doubling the time of anesthesia exposure produces a 17 percentage point increase in the probability that patients will underachieve. Cleft lip reduces while cleft right-left increases the duration of anesthesia exposure relative to cleft lip palate. Results do not change when anesthesia exposure only up to 4 years and socioeconomic factors are considered.

**Conclusions:** Both exposure to anesthesia and different types of orofacial cleft may result in underachievement at the CEB exam.

## Introduction

Orofacial clefts are one of the most frequent neonatal congenital malformations requiring corrective surgical procedures often since birth and extending sometimes to adolescence.^1^ A review of 67 studies from 19 countries during 1990-2017 identifying all measures used to assess long-term neurocognitive outcomes following general anesthesia and surgery in children up to the age of 18 concludes that studies vary significantly across important characteristics: the study population, surgery performed with possible confounding comorbidities, age at anesthesia exposure, follow-up, indication for and type of surgery, and outcomes.^2^ A review of psychological, behavioral, neuropsychological and academic outcomes of patients with cleft lip and/or palate from infancy to young adulthood shows a high incidence of reading problems and learning disabilities at school age, lower college attendance, and abnormal neural blood flow in young male adults similar to those observed in cases of dyslexia.^3^ Children with clefts score significantly lower than controls on reading and phonological memory,^4^ and have poorer academic achievement in comparison with the general population in Swedish^5^ and UK studies.^6^

Research assessing simultaneously the effects of anesthesia and oral cleft types on academic achievement is scarce, however. Available results are contradictory with some attributing impaired memory and learning later in life to anesthesia^7^ and others to the oral cleft type.^8^ One possible reason for contradictory results is the use of statistical methodologies that do not control for the complex interactions among comorbidities, the oral cleft type and the cumulative duration of anesthesia exposure.^2 3^ This study contributes to the literature investigating whether there is an association between anesthesia received during the treatment of orofacial clefts since birth and later academic achievement, distinguishing such association from the possible association between orofacial cleft types and academic achievement.

## Methods

### Study Population

Patients with orofacial clefts participating in this research are recorded in the database of the Centre Labio-palatin Albert de Coninck (Cliniques Universitaires Saint-Luc).^9^ The population comprised 110 patients (2017-2018), of which 88 could be contacted and received a detailed survey approved before hand by the ethics committee. Only 50 patients met the criteria of having been born between 1995 and 2007 and of not suffering from a syndromic form of orofacial cleft. Analysis could be done on the cohort of 29 patients for whom all academic scores of the Certificat d’Études de Base (CEB) could be confidently obtained. For the remaining 21, exam scores could not be obtained (4 cases), the exam was never completed because children failed twice (2 cases), they had not passed the exam yet (5 cases), exam scores of the general population were not published (5 cases), or exam scores were not yet available (5 cases). The 29 patients attended regular schools and underwent their academic evaluation in one of the following years: 2007, 2008, 2010, 2011, 2012, 2013, 2014, and 2016. There are no data for years 2009 and 2015. See table 1.

**Table 1:**
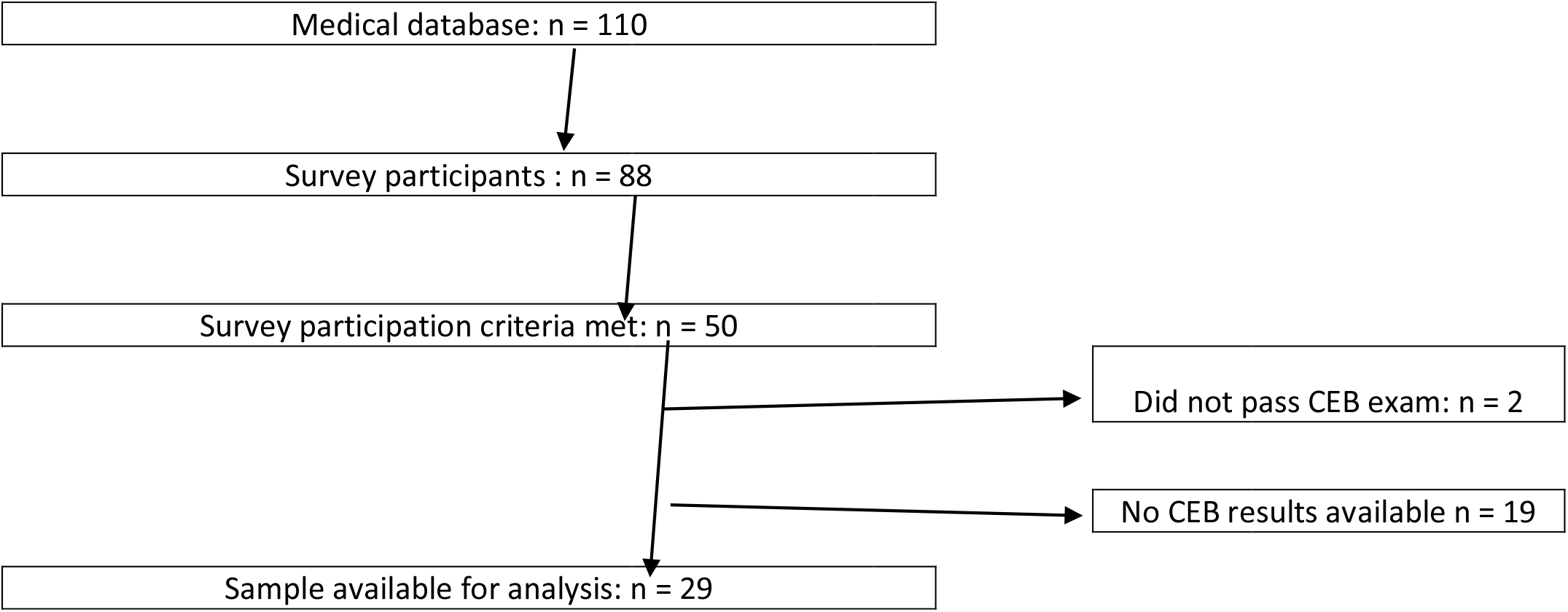
Flow chart of the study.

### Epidemiology Assessment

Patients were given a detailed survey asking information about their sex, age, country of origin, first spoken language, community, orofacial cleft type, number of operations, date of diagnosis, child’s and family medical history, number of children and place taken in the family, family income, mother and father final education level achieved, year the child passed the CEB exam and score obtained, number of times the child repeated a grade, school type attended, and extracurricular activities (Table 2). The data on the duration of anesthesia exposure, complications and desaturation during the operations were obtained from the Centre Labio-palatin Albert de Coninck’s database. Survey responses were cross checked with records from this database. An approval by the ethics committee (“Comité d’Ethique hospital-facultaire”) of the Catholic University of Louvain was given prior to the study. (ID: B403201525304).

**Table 2:** Sample statistics.

| Variable | Children With Oral Clefts n=50<br>n=29 | Variable | Children With Oral Clefts n=50<br>n=29 |
| --- | --- | --- | --- |
| **Oral clefts** |  | **Family medical history** |  |
| CL | 10,3% | yes | 20,7% |
| CP | 10,3% | no | 79,3% |
| CLP | 79,3% | **Family income (euro)** |  |
| **Child's gender** |  | <20000 | 10,3% |
| Female | 31,0% | 20000-40000 | 55,2% |
| Male | 69,0% | 40000-60000 | 27,6% |
| **Married mother** |  | >60000 | 6,9% |
| yes | 58,6% | Not available | 0,0% |
| no | 41,4% | **Diagnosis in utero** |  |
| **Mother : does she smoke** |  | Yes | 44,8% |
| yes | 6,9% | No | 55,2% |
| no | 93,1% | **Full term pregnancy** |  |
| **Mother's alcohol/drug consumption** |  | Yes | 10,3% |
| yes | 10,3% | No | 89,7% |
| no | 89,7% | **Extracurricular activities** |  |
| **Complications during pregnancy** |  | Yes | 62,1% |
| yes | 37,9% | No | 37,9% |
| no | 62,1% | **Adopted** |  |
| **Father : does he smoke** |  | Yes | 0,0% |
| yes | 27,6% | No | 100,0% |
| no | 72,4% | **Other medical problems** |  |
| **Place taken within the family** |  | Otitis | 79,0% |
| First | 44,8% | Phonetic | 58,0% |
| Second | 41,4% | Allergies | 34,0% |
| Third | 13,8% | **Average times repeated** | 0,5 |
| Fourth | 0,0% | **Average number of children** | 2,6 |

### Description of the ‘Certificat d’Études de Base’

The CEB is the standardized academic examination undergone by all students in the Belgian French community at the end of the ordinary primary school (11 years old) to qualify for entering middle school.^10^ The examination is also possible for students in the specialized primary and middle schools upon decision of the Class Board as well as for all minors of at least 11 years of age as of December 31^st^ upon parents’ approval. The Belgian population scores are the averages of children who attended all schooling program types. Children attending ordinary school programs represent almost 90% of the respective year population, which varies between 52,000 and 57,000 students. Pupils are tested on French, Mathematics, History, Geography and Science and must obtain at least 50% in each subject to pass the exam.

### Variables Used for Correlation and Regression Analyses

Academic achievement is measured as the ratio of the patients’ score in the CEB exam to the average general population’s score in the same year in logarithms (LPRO). This achievement measure is superior to the subjective teacher’s assessment^6^ and is robust to the criticism that the cumulative lifetime effect of a small impairment in children’s recollection memory may be substantially more apparent than at an earlier age.^7^ To tighten the achievement measure further, a categorical score variable (BEL) indicating whether the patient’s score is below the 25% percentile of the year’s population score is used given that this threshold normally identifies children in need of academic assistance.^11^ As the Swedish study^5^ and the UK^6^ study, this research uses the general population as control.

The duration anesthesia exposure (DUR) is measured as the minutes elapsed between induction and the end of anesthesia during all operations undergone by patients until the CEB examination date. This study uses several categorical variables: prenatal diagnostic of orofacial cleft anomaly (PRE), premature birth (PRM) (< 37 weeks of gestation), allergies (ALL), cleft lip (CL), cleft palate (CP), cleft lip-palate (CLP), bilateral orofacial cleft (CRL), phonetic problems (PHO), otitis (OTI), complications during operation (COM), low oxygen saturation defined as a saturation level during operations at or below the 95% as measured by pulse oximetry (DES), father’s highest education level (FED), mother’s highest education level (MED), extra-curricular activities (EXT), siblings (SIB), complications during pregnancy (PGY), alcohol and/or drugs consumption during pregnancy (DRU). Categorical variables were constructed by attributing a 1 to the illness or situation hypothesized to reduce academic achievement, and 0 otherwise.

### Statistical Analyses

First, correlation analysis is used to illustrate that the complex set of significant correlations among comorbidities and between them and duration of anesthesia exposure reported in the literature^3^ is also present in this study’s sample. This complexity together with the fact that the orofacial cleft type affects the duration of anesthesia exposure and the age of the first exposure (Table 3), make it necessary to use multivariate regression analysis.

**Table 3:**
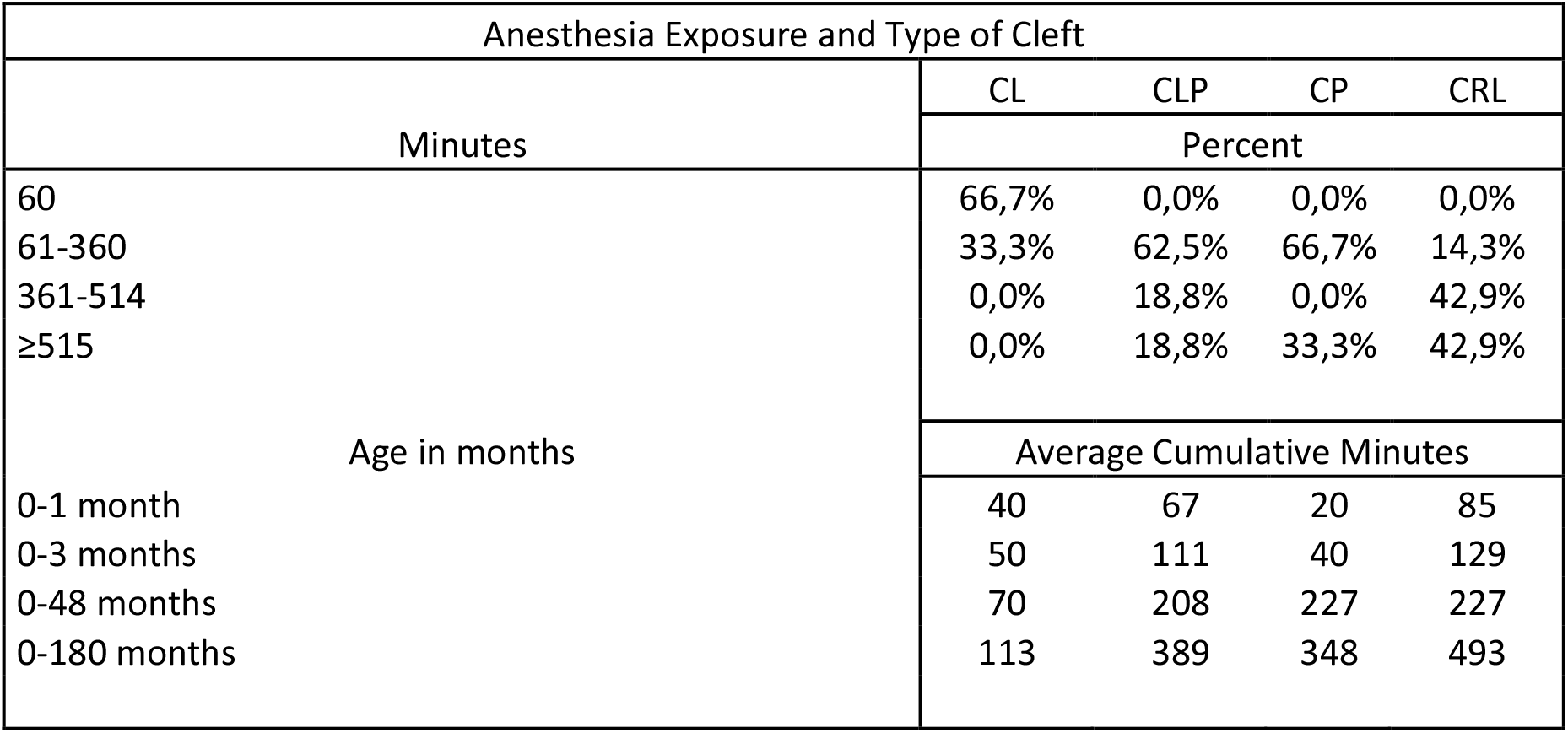
Anesthesia Exposure and Type of Cleft.

Second, given that controls used in the regressions and cleft types and anesthesia exposure are correlated among themselves, standard regression analysis will *produce biased and inconsistent coefficient estimates*, even if the sample size goes to infinity.^12^ While recognized, these interactions have not necessarily led to use appropriate statistical methodologies.^7 8^ This motivates the simultaneous multivariate regression methodology used (see Technical Appendix). The software was RATS version 9.2. Statistical significance is at P-values equal to or lower than 0.10.

## Results

### Correlation Analysis

Pearson’s correlation analysis shows the complex set of significant correlations between comorbidities and socioeconomic factors, and between them and DUR. DUR, PRE, CL, PHO are the main variables correlated with children’s achievement (measured as BEL) or LAGD (Table 4). For instance, CLP and COM are positively correlated with a p-value of 0.01. While CL is negatively correlated with DUR with a p-value of 0.01, CRL is positively correlated with DUR with a p-value of 0.05. Although illnesses display in general low correlations with achievement measured as LPRO, DUR and BEL are positively correlated with a p-value of 0.05. Finally, socioeconomic variables are significantly correlated among themselves (e.g. INC and MED) and with medical variables (e.g. EXT and CLP).

**Table 4:** Correlation analysis.

| T-test for correlation coefficients |  |  |  |  |  |  |  |  |  |  |  |  |  |  |  |  |  |  |  |  |  |  |  |  |  |
| --- | --- | --- | --- | --- | --- | --- | --- | --- | --- | --- | --- | --- | --- | --- | --- | --- | --- | --- | --- | --- | --- | --- | --- | --- | --- |
|  | LPRO | OPE | BEL | PRE | PRM | ALL | INC | SEX | CL | CP | CLP | CRL | PHO | OTI | COM | DES | DUR | FED | MED | EXT | SIB | PGY | DRU | LAGD |  |
| LPRO | N.A. |  |  |  |  |  |  |  |  |  |  |  |  |  |  |  |  |  |  |  |  |  |  |  | LPRO |
| OPE | -1.85 | N.A. |  |  |  |  |  |  |  |  |  |  |  |  |  |  |  |  |  |  |  |  |  |  | OPE |
| BEL | -6.67 | 1.09 | N.A. |  |  |  |  |  |  |  |  |  |  |  |  |  |  |  |  |  |  |  |  |  | BEL |
| PRE | 0.98 | -2.37 | -0.73 | N.A. |  |  |  |  |  |  |  |  |  |  |  |  |  |  |  |  |  |  |  |  | PRE |
| PRM | 0.23 | -0.74 | 0.38 | 0.41 | N.A. |  |  |  |  |  |  |  |  |  |  |  |  |  |  |  |  |  |  |  | PRM |
| ALL | -1.38 | -0.49 | 0.52 | 0.37 | -0.04 | N.A. |  |  |  |  |  |  |  |  |  |  |  |  |  |  |  |  |  |  | ALL |
| INC | -0.61 | 0.95 | 0.61 | -0.70 | -1.52 | 1.75 | N.A. |  |  |  |  |  |  |  |  |  |  |  |  |  |  |  |  |  | INC |
| SEX | 0.19 | -0.81 | -0.06 | 0.33 | -1.62 | -0.20 | 1.23 | N.A. |  |  |  |  |  |  |  |  |  |  |  |  |  |  |  |  | SEX |
| CL | 0.16 | -2.50 | -1.01 | 0.41 | -0.60 | 1.23 | -0.67 | -1.62 | N.A. |  |  |  |  |  |  |  |  |  |  |  |  |  |  |  | CL |
| CP | -1.33 | 1.03 | 0.38 | 0.41 | 1.38 | 1.23 | 0.95 | -0.23 | -0.60 | N.A. |  |  |  |  |  |  |  |  |  |  |  |  |  |  | CP |
| CLP | 0.39 | -0.02 | 0.97 | 0.12 | 0.41 | -1.18 | -0.20 | 0.33 | -2.11 | -2.11 | N.A. |  |  |  |  |  |  |  |  |  |  |  |  |  | CLP |
| CRL | 0.35 | 0.92 | -0.68 | -0.73 | -1.01 | -0.37 | 0.04 | 0.88 | -1.01 | -1.01 | -4.17 | N.A. |  |  |  |  |  |  |  |  |  |  |  |  | CRL |
| PHO | -0.31 | 2.09 | 0.77 | -1.85 | 1.55 | -1.48 | 0.65 | -0.25 | -2.30 | 1.55 | -1.03 | 1.70 | N.A. |  |  |  |  |  |  |  |  |  |  |  | PHO |
| OTI | -0.03 | 0.78 | 0.47 | 0.28 | 0.92 | -0.88 | -0.21 | -0.65 | -2.17 | 0.92 | 0.28 | 0.47 | 2.51 | N.A. |  |  |  |  |  |  |  |  |  |  | OTI |
| COM | 0.35 | 0.63 | 0.09 | -1.22 | -0.29 | -0.88 | -0.65 | -0.56 | -1.55 | -1.55 | 2.81 | -0.77 | 0.72 | 1.38 | N.A. |  |  |  |  |  |  |  |  |  | COM |
| DES | 1.80 | 0.42 | -0.12 | -0.12 | 0.78 | -2.02 | 0.20 | -0.33 | -1.67 | -1.67 | 2.23 | -0.12 | 0.28 | -0.28 | 1.22 | N.A. |  |  |  |  |  |  |  |  | DES |
| DUR | -2.04 | 8.29 | 2.06 | -3.52 | -0.47 | -0.87 | 0.77 | -0.64 | -3.42 | -0.36 | 0.27 | 2.14 | 2.51 | 0.99 | 1.24 | 1.32 | N.A. |  |  |  |  |  |  |  | DUR |
| FED | -0.69 | 1.26 | 0.57 | 0.05 | 0.17 | -2.84 | -2.83 | -0.03 | -1.07 | -2.51 | 1.61 | 0.57 | 0.34 | 0.67 | 1.19 | -0.05 | 1.99 | N.A. |  |  |  |  |  |  | FED |
| MED | -0.16 | 0.46 | -0.09 | 1.22 | 0.29 | -1.48 | -2.58 | -2.02 | 0.29 | -0.92 | 1.22 | -0.95 | -0.72 | 1.41 | 0.72 | -0.46 | 0.01 | 2.98 | N.A. |  |  |  |  |  | MED |
| EXT | -0.54 | 1.74 | 1.19 | -0.05 | -0.17 | 0.16 | 1.00 | -1.71 | -1.43 | -1.43 | 2.40 | -0.57 | 0.41 | 1.19 | 1.11 | 0.80 | 1.79 | 0.91 | 1.19 | N.A. |  |  |  |  | EXT |
| SIB | -0.08 | -0.54 | 0.04 | 0.84 | 1.02 | 0.68 | -1.55 | 0.12 | 1.02 | -0.71 | -0.22 | 0.04 | -0.36 | -1.57 | 0.36 | -0.84 | -0.27 | 1.71 | 0.70 | -1.71 | N.A. |  |  |  | SIB |
| PGY | -0.13 | 0.15 | 0.30 | -0.80 | 1.07 | -0.62 | -0.02 | 1.78 | -1.43 | 2.51 | -0.80 | 0.30 | 1.19 | 1.19 | -0.41 | -1.49 | 0.09 | -0.63 | -1.11 | -2.73 | 0.52 | N.A. |  |  | PGY |
| DRU | 0.21 | 0.49 | -0.23 | 0.23 | 0.76 | 0.28 | 0.40 | 1.53 | -0.82 | 2.58 | 0.23 | -1.38 | 1.05 | 0.04 | -0.07 | -0.23 | -0.27 | -1.10 | 0.07 | -1.98 | 1.92 | 1.10 | N.A. |  | DRU |
| LAGD | -1.45 | 4.56 | 1.34 | -3.28 | 0.60 | -0.50 | 1.44 | -1.27 | -3.22 | 0.56 | 0.55 | 0.93 | 2.97 | 1.41 | 1.49 | 1.05 | 6.75 | 0.49 | 0.48 | 1.89 | -1.08 | 0.69 | 0.22 | N.A. | LAGD |
|  | LPRO | OPE | BEL | PRE | PRM | ALL | INC | SEX | CL | CP | CLP | CRL | PHO | OTI | COM | DES | DUR | FED | MED | EXT | SIB | PGY | DRU | LAGD |  |
| T-test for Ho: rho=0, with 2.05 and 2.77 as significant levels at 5% (in bold) and 1% (circled), respectively. T-tests are adjusted to account for the non-normality of some variables. |  |  |  |  |  |  |  |  |  |  |  |  |  |  |  |  |  |  |  |  |  |  |  |  |  |

### Regression Analysis

Table 5, panel a, displays the regression results for the dependent variables BEL and LDUR. A 100% increase in LDUR generates a 17 percentage point increase (p-value = 0.06) in the probability that patients will underachieve at the CEB exam. The orofacial cleft type has a direct impact on LDUR. LDUR increases 38% with CRL (p-value = 0.00) and decreases 75% with CL (p-value = 0.00), relative to CLP. SEX is the only significant epidemiological factor affecting LDUR beyond the cleft type. Male patients have a 28% reduction in LDUR (p-value = 0.01) relative to female patients as the average duration of anesthesia exposure of male patients is 12% lower than female patients. Socioeconomic conditions such as INC and FED, when statistically significant, do not alter the results. The only significant confounder is FED: father university education increases LDUR 32% (p-value = 0.01).

Evidence that the possible negative effects of anesthesia are more significant at early age (i.e. up to 48 months of age)^11^ is scarce.^2^ Table 5, panel b, displays the regression results for the dependent variables BEL and LAGD. Exposure to anesthesia up to the age of 48 months increases the probability of underachievement 23 percentage points (p-value = 0.07). The medical factors explaining LAGD are consistent with those of the upper panel a, except that CRL and FED are not statistically significant. The insignificance of CRL is consistent with the fact that this orofacial cleft type is usually subject to operation later in life (see Table 3). FED insignificance may suggest that education and health care effects accumulate over time.

**Table 5:** Regression Results.

| a. Dependent Variable: BEL |  |  |
| --- | --- | --- |
|  | CONSTANT | LDUR |
| Coefficient | -0,72 | 0,17 |
| P-value | 0,13 | 0,06 |

| Dependent Variable: LDUR |  |  |  |  |  |  |
| --- | --- | --- | --- | --- | --- | --- |
|  | CONSTANT | SEX | CL | CP | CRL | FED |
| Coefficient | 5,95 | -0,33 | -1,40 | -0,08 | 0,32 | 0,28 |
| P-value | 0,00 | 0,01 | 0,00 | 0,83 | 0,00 | 0,01 |

| b. Dependent Variable: BEL |  |  |  |  |  |  |
| --- | --- | --- | --- | --- | --- | --- |
|  |  | CONSTANT | LAGD |  |  |  |
|  | Coefficient | -0,93 | 0,23 |  |  |  |
|  | P-value | 0,13 | 0,07 |  |  |  |
| Dependent Variable: LAGD |  |  |  |  |  |  |
|  | CONSTANT | SEX | CL | CP | CRL | FED |
| Coefficient | 5,55 | -0,37 | -1,20 | 0,04 | 0,11 | 0,02 |
| P-value | 0,00 | 0,00 | 0,00 | 0,89 | 0,37 | 0,87 |

If the dependent variable is in logarithms, the effect of a categorical variable on it is the exponential of the coefficient value minus 1.
Estimates are tested and corrected if needed for heteroscedasticity and serial correlation in the residuals.
$\chi^2(2)$ test that all regressors are zero in first equation = 9.95, p-value = 0.01 and with FED 9.96, p-value 0.01.
$\chi^2(6)$ test that all regressors are zero in second equation = 11409.73, p-value = 0.01 and with FED $\chi^2(6)$ = 11508.61 p-value 0.00.
Cumulated periodogram test for absence of serial correlation in each equation, maximum gaps = 0.23 and 0.12, respectively, for an approximate rejection limit of 0.30 at 10% confidence level. With FED, maximum gap = 0.23 and 0.12, respectively.

## Discussion

Most studies have assessed the effects of anesthesia in children without a congenital malformation undergoing one elective surgical procedure or one anesthesia exposure of less than 60 minutes.^14 15^ School underachievement following an early single short exposure to anesthesia has also been reported.^16-18 7^ This study finds possible CEB exam underachievement following an average cumulative anesthesia exposure of 382 minutes and is thus consistent with research finding a negative effect of anesthesia on achievement after long or several exposures.^19^

Lack of data on the timeliness of cleft surgical repair^20^ and the duration of anesthesia exposure is frequent in studies suggesting either underachievement in children with orofacial clefts^2 13^ or the absence of negative effects.^21^ Consistent with the few studies controlling for the age at anesthesia exposure,^11^ a strength of this study is to show that a cumulative average exposure of 207 minutes up to 48 months of age is already associated with underachievement.

Comparisons across studies are difficult by the lack of a standard classification of orofacial cleft types. Some studies distinguish between CL, CP and unilateral and bilateral CLP^6^, or CL, CP and CLP^5^, while this study also includes CRL. However, the consistency of larger effects across cleft types involving the palate than those only involving the lip is reassuring.

Comparisons are also difficult because of differences regarding the measurement of anesthesia exposure duration. An additional strength of this study is to measure anesthesia exposure as the minutes elapsed between induction and end of anesthesia administration. Studies often use the total time spent by the patient in the operating room, which may bias results as the number of operations increases.^7^

The low power of small samples in anesthesia-related neurotoxicity research is pervasive, with samples as low as 15 or 21.^2^ The size of the current study, 29, is close to the statistical convention of 30 observations for parametric tests, and statistics are corrected for sample size. An important additional contribution of this study is its statistical methodology, which avoids obtaining biased and unstable coefficients estimates such as in the Danish study.^8^

Comorbidities (e.g. OTI, PHO, a combination of both) or COM were statistically insignificant (results not shown). Socioeconomic factors did not affect school achievement, except FED when anesthesia exposure up to the CEB exam was considered. This may be due to the known fact that higher educated individuals spend more in medical care and education, which beneficial effects on children’s development accumulate over time. In contrast to the American study,^13^ for example, most parents in this study had high-school education.

There are some limitations. First, as the Swedish^5^ and UK^6^ studies using the general population as control, this research lacks comparative data concerning anesthesia exposure for a matched population of children *without* orofacial cleft malformations. This limits a thorough assessment of medical and socioeconomic variables effects on school achievement. Second, the Belgian children general population includes those with orofacial malformations and special education schooling. While their share is small, it might still produce a bias in the results, albeit making it more difficult to find the significant effects obtained. Finally, the current study does not control for the improved academic achievement of children born with orofacial clefts over time partly because of surgical techniques’ advances and the implementation of a multidisciplinary approach.^6^ However, a systematic multidisciplinary approach is part of the standard operating procedures of the Centre Labio Palatin Albert Coninck, which may produce a *downward bias* in the estimated negative impact of anesthesia on achievement.

## Conclusions

Exposure to anesthesia by children born with an orofacial cleft may result in underachievement during the CEB exam, and different forms of orofacial clefts can also affect achievement indirectly. While patients benefit from early operations that help their overall development including speech, hearing, learning and even nourishment, these benefits must be balanced with the possible negative effects of anesthesia on academic achievement.

## Data Availability

Data is available upon request

## Contributorship and Funding

Dr. Bénédicte Bayet and Prof. Dr. Francis Veyckmans are thanked for scientific guidance and Dr. Gaston Giordana and Prof. Francisco Nadal De Simone for statistical guidance. The author declares having no conflict of interest and not having received financial support for this study.

**Abbreviations**
AGD: Minutes of anesthesia exposure up to 48 months of age ALL: Allergies
BEL: Patients with a score below the 25% percentile of the same year Belgian population’s average score
CEB: Certificat d’Études de Base
CL: Cleft lip
CLP: Cleft lip-palate
COM: Complications during operations
CP: Cleft palate
CRL: Cleft right-left
DES: Low oxygen saturation
DRU: Alcohol and/or drugs consumption during pregnancy
DUR: Minutes of anesthesia exposure until the CEB exam
EXT: Extra-curricular activities
FED: Father’ highest education level
INC: Family income in logarithms
LDUR: Logarithm of DUR
LGAD: Logarithm of AGD
LPRO: Logarithm of the ratio of the CEB patients’ score to the population’s average score in the same year
MED: Mother’s highest education level
OPE: Number of operations underwent by the patient
OTI: Otitis
PGY: Complications during pregnancy
PHO: Phonetic problems
PRE: Prenatal diagnostic of orofacial cleft anomaly
PRM: Premature birth
SEX: Sex of the patient
SIB: Siblings

## Notes

### Competing Interest Statement

The authors have declared no competing interest.

### Funding Statement

This study has not received any funding

### Author Declarations

An approval by the ethics committee (Comite d Ethique hospital-facultaire) of the Catholic University of Louvain was given prior to the study. (ID: B403201525304).

